# Time-adjusted Analysis Shows Weak Associations Between BCG Vaccination Policy and COVID-19 Disease Progression

**DOI:** 10.1101/2020.05.01.20087809

**Authors:** Katarína Bod’ová, Vladimír Boža, Broňa Brejová, Richard Kollár, Katarína Mikušová, Tomáš Vinař

## Abstract

In this study, we ascertain the associations between BCG vaccination policies and progression of COVID-19 through analysis of various time-adjusted indicators either directly extracted from the incidence and death reports, or estimated as parameters of disease progression models. We observe weak correlation between BCG vaccination status and indicators related to disease reproduction characteristics. We did not find any associations with case fatality rates (CFR), but the differences in CFR estimates are at present likely dominated by differences in testing and case reporting between countries.

## Introduction

The reports on a possible use of the well-established and widely used BCG vaccine as a protection against COVID-19 (de Vrieze, 2020) raised a lot of interest and media coverage. Currently, four clinical trials have been designed to evaluate the potential of BCG for protection against the SARS-CoV-2 infection in health-care workers (Bonten, 2020; Khattab, 2020; Curtis, 2020; Cirillo and DiNardo, 2020). These studies are driven by the so called non-specific effects of BCG vaccine on viral infections, observed in animal models, as well as in humans, although the molecular basis of this phenomenon is not completely understood (Moorlag et al., 2019).

The associations between BCG vaccination policy and COVID-19 disease progression have also been a subject to controversy in data analysis, with some studies claiming significant effects on the number of cases and case fatality rates (Miller et al., 2020; Berg et al., 2020), while others criticizing weaknesses of those studies and claiming no statistically significant differences (Szigeti et al., 2020; Hensel et al., 2020; Fukui et al., 2020; Singh, 2020).

While correcting for many covariate factors (such as population size, population age distribution, etc.), most of these studies, however, failed to correct for the differences in time progression of the epidemics in each country. COVID-19 epidemic usually starts from relatively few imported cases and spreads quickly through exponential growth with high reproduction numbers. At unchecked growth rates, a significant percentage of the country population would be infected before the disease would subside. However, this growth rate only continues until effective measures, such as lockouts or social distancing policies, are introduced, changing the dynamics of the epidemics substantially, with infection rates rarely reaching a significant percentage of the whole population in the first wave Flaxman et al. (2020). In this study, we have estimated a variety of indicators characteristic for different stages of COVID-19 epidemics, also adjusting for time since the beginning of the epidemics in each country, and found that several key indicators show weak, but statistically significant, associations with BCG vaccination status.

## Results

To compare the COVID-19 disease progression between countries with recent universal BCG vaccination policy and those without, several parameters derived from the case and death reports in each country were selected. The parameters reflect early-stage disease spread characteristics (when they are likely not yet affected by social distancing policies), early-stage case fatality rates (before potential effects from overwhelmed health care system), and progression of the disease after the changes characteristic for social distancing policies take effect.

### Estimates of early stage *R* are lower in countries with recent BCG vaccination policies

The reproduction number *R*, the average number of secondary cases of disease caused by a single infected individual, has been estimated using EpiEstim package (Cori et al., 2013), based on 7-day windows, the first estimate starting on the day when cumulative number of 100 reported cases have been reached (R100), the second estimate starting on 10th day afterwards (R100+10). In many countries, this time period would not reflect the effects of social distancing policies, but would also somewhat avoid the initial period when the case reporting is likely to be unreliable. In both cases, the countries with recent BCG vaccination policies show lower *R* estimates (Figure 1) and these shifts were statistically significant (Mann Whitney U-test, *P* = 0.04 for R100 and *P* = 0.006 for R100+10).

**Figure 1:**
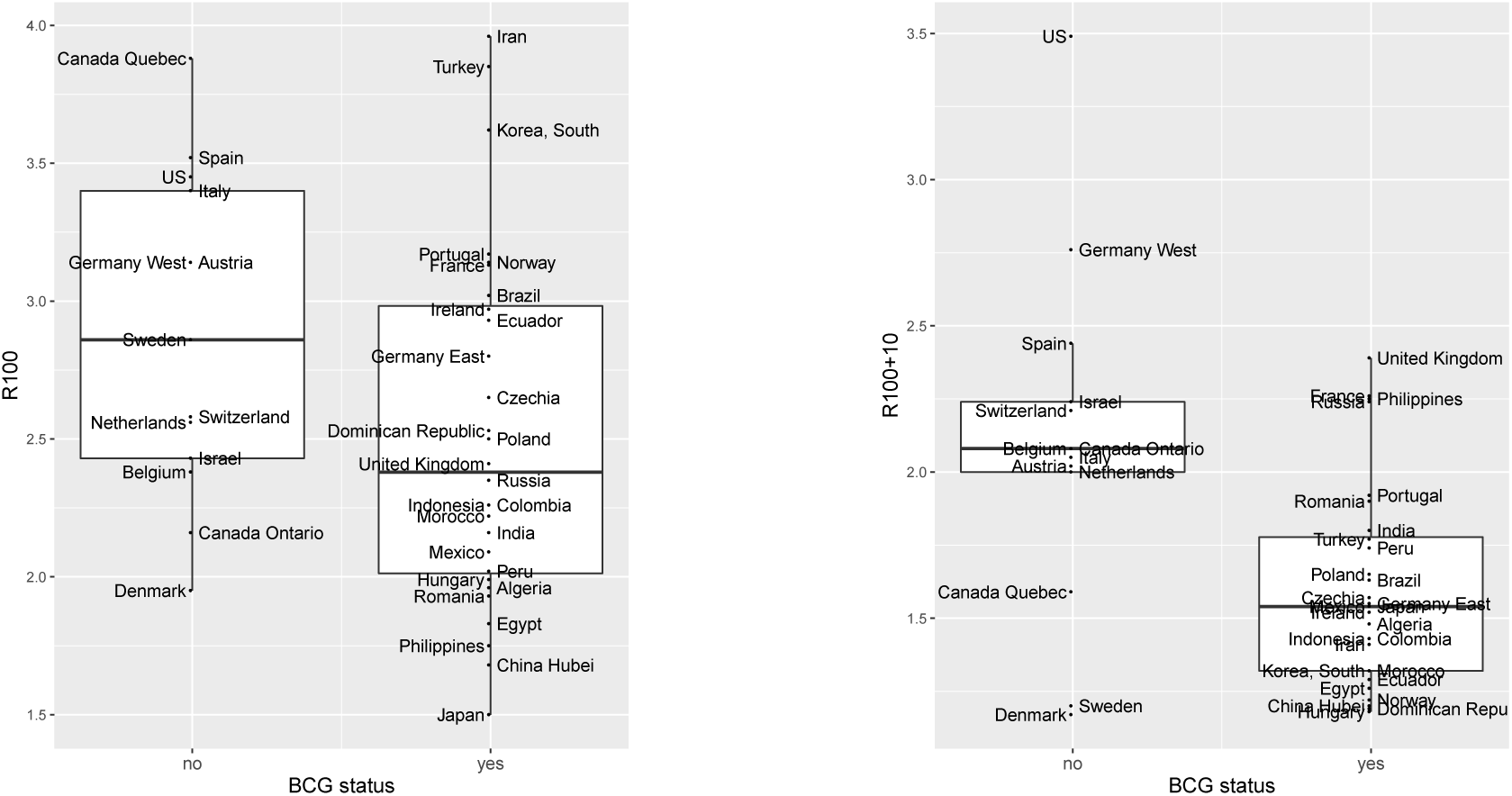
Comparison of estimated reproduction numbers R100 (left) and R100+10 (right) between countries with and without the universal BCG vaccination policy.

We have also examined the number of days between 10 and 100 reported cases (C10), 100 and 1000 reported cases (C100), 10 and 100 reported deaths (D10), and 100 and 1000 reported deaths (D100). These time periods reflect R in various early stages of the epidemic, longer periods meaning slower spread of the disease. Note, that C10 numbers are likely unreliable (due to initial problems in establishing testing and reporting policies in each country), and there are only few countries that reached 1000 reported deaths before our data set cutoff. Also note that if we assume a constant case fatality rate within a specific time period (typically 6-10 days) and a specific country, and also assume exponential growth in cases within this time period, the numbers D10 and D100 do not actually reflect the death rate, but instead only depend on the underlying value of R. Death reports are likely more accurate than case reports, which are much more affected by testing and reporting policies in each country. On average, all of these time periods are slightly longer in countries with recent universal BCG policies, with statistically significant results for D10 (Mann-Whitney U-test, *P* = 0.02).

### No differences in case fatality rates

We have estimated case fatality rates on days when 100 and 1000 cumulative deaths were first reached in each country (CFR100 and CFR1000 respectively), and also used CMMID methodology (Nishiura et al., 2009; Russel et al., 2020) to correct for estimation of active cases (cCFR100 and cCFR1000). While some small shifts were observed between countries with and without recent universal BCG vaccination policies (see Supplementary material), these shifts are not statistically significant.

### Significant differences in the coefficients of the Vazquez model

One of the difficulties in modelling and predicting the extent of the coronavirus spreading in a population is the divergence of the observed data (the number of confirmed active cases in individual countries) from the trends expected from the traditional SIR type models. Ziff and Ziff (2020) have recently observed that deaths in China do not follow the typical epidemiological curve and instead of an exponential growth they follow a combined polynomial growth with exponential decay (PGED). Polynomial growth has been also confirmed for multiple other countries (Merrin, 2020) and even though the initial spread in many countries is approximately exponential, it is followed by a steady polynomial growth and in a longer run by an exponential decay (Komarova and Wodarz, 2020).

For a possible explanation of the transition from exponential to polynomial growth, it is natural to look into self-imposed or government-imposed social distancing measures. These measures transform the structure of virus transmitting contact networks in a population, possibly to small-world network structures or even fractal networks. In contrast to small-world networks, social networks under standard conditions contain a significant fraction of nodes with high number of connections (that correspond to potential superspreaders). Interestingly, polynomial growth of the number of infections in time in well connected scale-free networks emerges naturally as a consequence of infection initially reaching the highly connected nodes and their neighbors, while their isolation or recovery significantly reduces the interconnectivity of the residual network (Szabó, 2020). Theoretical study of the infection spread in scale-free networks by Vazquez (2006) leads to an explicit formula for the number of infected individuals in time in a form of PGED. The formula contains three key parameters: *p* - the coefficient of the polynomial growth (not necessary an integer), *τ* - the rate of decay of the exponential tail (1/*τ* is an analogue to the rate of removal of individuals from the infected class to inactive recovered class in the traditional SIR-type models), and *A* - the constant prefactor (scaling the total population). Based on the value of these parameters, it is straightforward to determine Nmax, the number of infected at the peak of the epidemic, which is independent of the choice of the reference time for the start of the infection. These parameters were obtained by the best fit on the linear scale to the data in each of the considered countries/regions.

Interestingly, we have found that the parameters *τ* and Nmax significantly differ between countries split into two groups—with and without recent universal BCG vaccination policies (Figure 2). The *τ* parameter shifts to the higher values, signifying higher recovery rate in the countries with recent universal BCG vaccination policies (Mann-Whitney U-test *P* = 0.04). In addition, these countries have generally lower numbers of infected cases at the peak of the epidemic (Nmax) corrected for underreporting (Mann-Whitney U-test *P* = 0.002).

**Figure 2:**
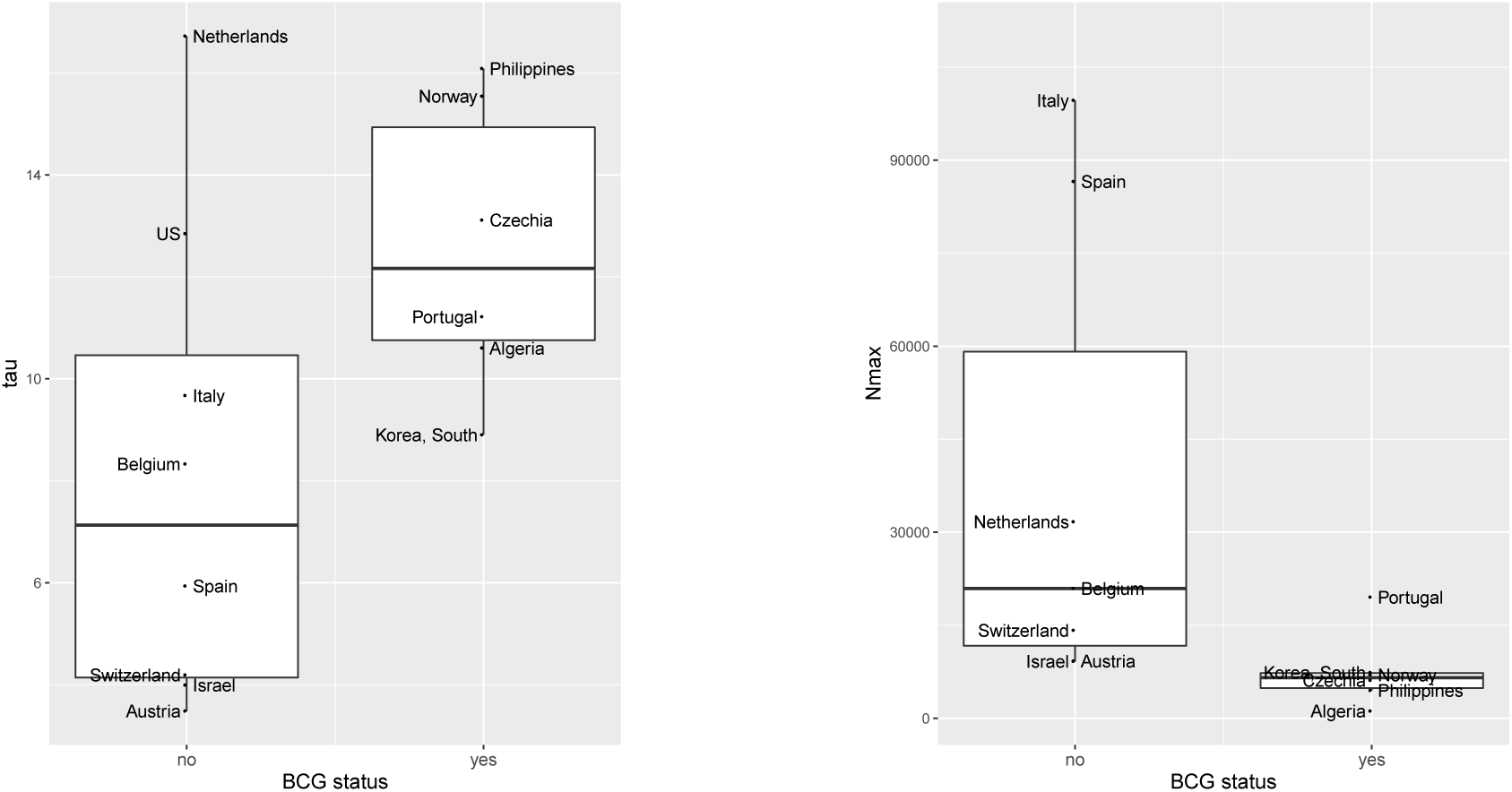
Comparison of Vazquez model parameters estimated for countries with and without universal BCG vaccination policies. Left: Rate of decay of the exponential tail (*τ*). Right: Number of infected cases at the peak of the epidemic corrected for underreporting (Nmax).

### East and West Germany

The case of Germany is interesting, since the country has been split into East and West Germany in 1949 and reunited in 1990. In East Germany, the policies regarding BCG vaccination followed Eastern Bloc practices, with universal vaccination policy in place between 1951 and 1998. In West Germany, the vaccination has been introduced in 1961, but in 1975 it was discontinued in favor of vaccinating high risk groups only. [The information has been reconstructed from the notes in BCG atlas, however we were not able to confirm this from other sources.] In the present crisis, the whole Germany follows similar practices in case reporting and treatment of the disease. Interestingly, East Germany exhibits much lower estimates of R than West Germany at the corresponding phases of the epidemic (R100 = 2.8, R100+10 = 1.55 in East Germany vs. R100 = 3.14, R100+10 = 2.76 in West Germany; see also Figure 3). Also, the death rate from COVID-19 seems to be significantly lower in East Germany, even when correcting for differences in age distribution (Table 1).

**Figure 3:**
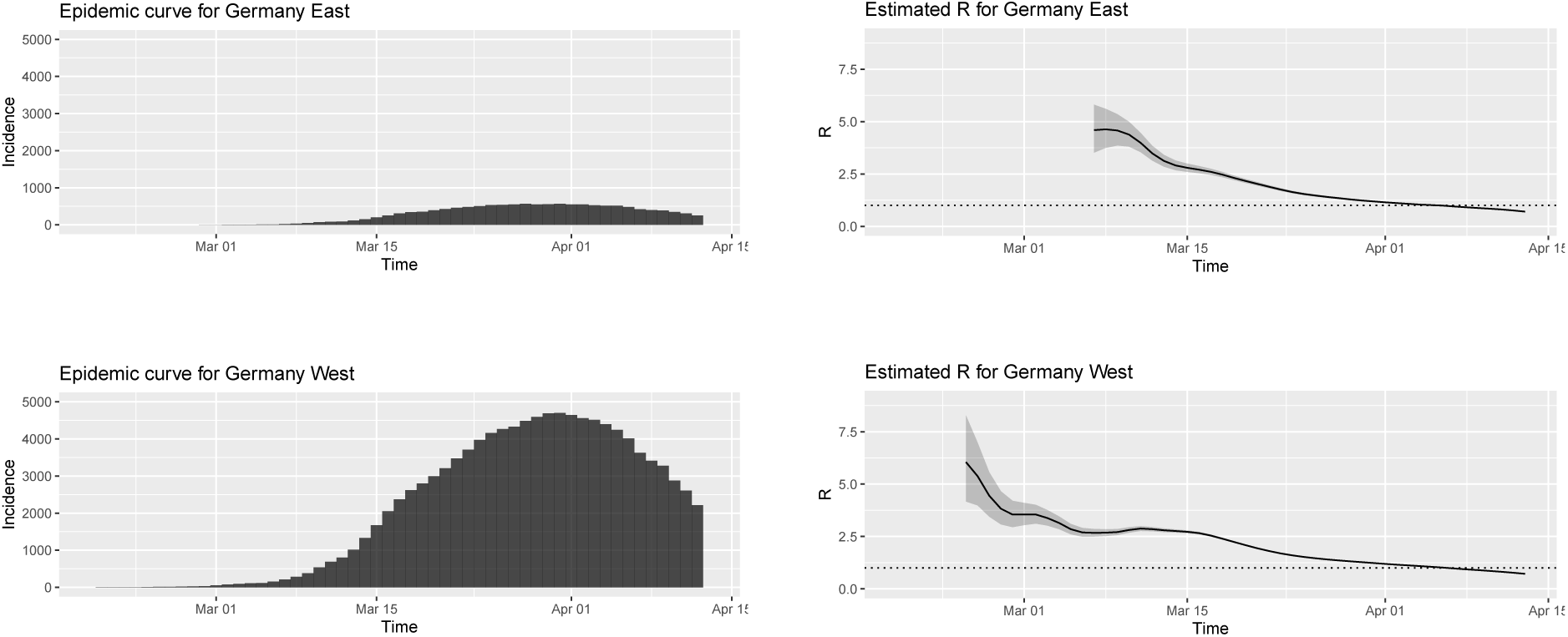
Comparison of COVID-19 epidemic progression between East Germany and West Germany. Reproduction numbers *R* were estimated using seven day windows using smoothed incidence numbers.

## Discussion

While some of the previous studies have observed associations between BCG vaccination policy and spread of COVID-19 (Miller et al., 2020; Berg et al., 2020), others criticized their work and showed that after corrections for various covariate factors, no statistically significant associations could be found Hensel et al. (2020); Fukui et al. (2020); Singh (2020). Most of these studies have used indicators that were quite straightforward, such as the number of reported cases per million inhabitants on a particular date. Here, we have instead chosen a variety of indicators that reflect characteristics of various phases of the epidemics in each country, and moreover, these indicators were implicitly or explicitly adjusted according to the time from the beginning of the epidemic in each country. In fact, we hypothesize that such time adjustment is one of the key factors in such an analysis considering what we know about the spread of COVID-19.

**Table 1:**
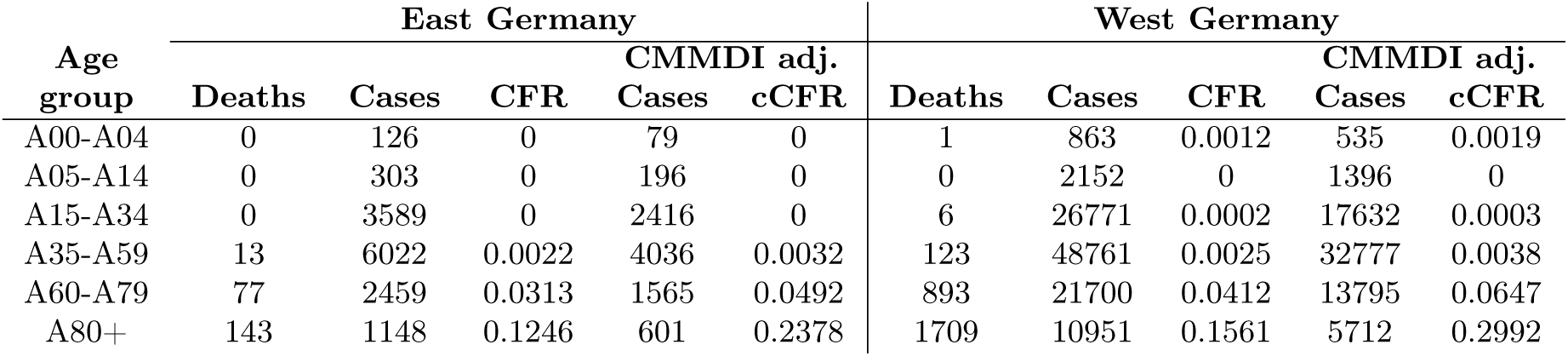
Differences in CFR in different age groups between East Germany and West Germany. Both raw CFR values and values corrected by CMMDI methodology are presented.

In our data, we have observed several statistically significant associations, and we conclude that there is an association between BCG vaccination policy and spread of COVID-19. However, whether this association is causal or is merely an observed correlation due to some other common factor, is impossible to say. Moreover, most observed shifts in various coefficients are rather small and while the universal BCG vaccination policy may have had a positive impact in some of the countries, the observed impact clearly cannot replace effective policies such as lockdowns and social distancing measures which currently constitute the most effective weapon against the epidemic. At best, the existence of universal BCG vaccination policy may have provided a few days time for governments to effectively institute such policies.

One of the interesting observations is that we did not find any correlation between BCG vaccination policy and CFR. While this may suggest a hypothesis that BCG vaccination may help to limit spread, but may not be effective against difficult progression of the disease in susceptible individuals, we would be careful to draw such conclusions. This is because the estimates of CFR are clearly unreliable at this point of time, with many countries showing CFR estimates well over 10%. Likely, huge differences between countries do not reflect real differences in outcomes of the disease, but rather discrepancies in the amount and effectiveness of testing, with many light or asymptomatic cases remaining undetected. In fact, such a conclusion is partly supported by the evidence from East/West Germany, where we can assume consistent reporting of cases and outcomes, and where differences in CFR seem to be consistent with historical differences in BCG vaccination policies, even after correcting for differences in the age distribution of the population.

## Methods

### Obtaining case and death reports

The information on reported cases, deaths, and recoveries related to COVID-19 assembled by John Hopkins University Center for System Science and Engineering (Dong et al., 2020) has been downloaded from Humanitarian Data Exchange (Humanitarian Data Exchange, 2020) on April 14, 2020. The data set covers reports from 266 countries from January 22, 2020 until April 13, 2020. For further analysis, only 41 countries with at least 100 reported cumulative deaths have been retained. We also used the data set for Germany maintained by Robert Koch Institute, containing reported cases, deaths, and recoveries split geographically and into age groups; the data set was downloaded through ArcGIS (Robert Koch-Institut and Bundesamt für Kartographie und Geodäsie, 2020). For our analysis, the data were split geographically into East Germany (Brande-burg, Mecklenburg-Vorpommern, Sachsen, Sachsen-Anhalt, Thüringen, and Berlin) and West Germany (Schleswig-Holstein, Hamburg, Niedersachsen, Bremen, Nordrhein-Westfalen, Hessen, Rheinland-Pfalz, Baden-Württemberg, Bayern, and Saarland).

### BCG status of individual countries

For countries included in the study, we have assembled information from the BCG World Atlas (Zwerling et al., 2011) and from the WHO-UNICEF estimates of BCG coverage (World Health Organization, 2020) (see Supplementary materials). Based on this information, the countries were divided into positive BCG status (the countries with current universal BCG vaccination policy and countries with past universal policies discontinued after 1990 or with recent reports of high vaccination coverage from WHO) and negative BCG status (the countries without universal BCG vaccination policy and those that discontinued universal BCG policies and did not satisfy the above conditions).

### Estimation and extraction of indicators

The indicators were extracted from the time series data sets using simple scripts, as outlined in the Results (see Supplementary Material for tables). All of the indicators are computed in time that is relative to a particular milestone, i.e. reaching a particular cumulative number of case reports or death reports. In this way, compared indicators are synchronized at a particular stage of the epidemic. Since the number of cases and deaths is highly dependent on the stage of the epidemic, using such synchronized indicators is a key in our analysis.

Case fatality rate indicators CFR100 and CFR1000 were computed on the days when the cumulative number of reported deaths surpassed 100 and 1000 respectively; the cumulative number of deaths was divided by the cumulative number of reported cases 7 days prior to that date. As alternative indicators for case fatality rates, denoted as cCFR100 and cCFR1000, we have used methodology established by the Centre for the Mathematical Modelling of Infectious Diseases (Nishiura et al., 2009; Russel et al., 2020), systematically compensating for confirmation-to-death delay using lognormal distribution with mean delay of 13 days and a standard deviation of 12.7 days (Linton et al., 2020). Regardless of the method, the main problem with CFR indicators is inconsistent reporting on the number of cases in different countries, as this depends highly on testing strategy, reporting methodology, as well as testing capacities of individual countries. Thus, CFR estimates are likely dominated by these factors. We are not aware of any simple method that could overcome this problem at this point of time.

Note that indicators D10 (time from 10 death reports to 100 death reports) and D100 (time from 100 death reports to 1000 death reports), even though based on the numbers of reported deaths, are unlikely to reflect CFR, but instead simply serve as more stable estimates reflecting the underlying reproductive number *R*. This is because if we assume exponential growth phase and a constant CFR over this period of time, the CFR coefficient will cancel out in the computation of the expected number of days to reach 10-fold increase in the number of deaths.

Indicators R100 and R100+10 were computed using EpiEstim R package (Cori et al., 2013). This method is based on Bayesian inference, modelling new infections as a Poisson process with rate governed by the instantaneous reproduction number and the number and total infectiousness of infected individuals at the current time interval. The instantaneous reproduction number has a gamma-distributed prior and during the inference is assumed to be constant within each seven-day sliding window to yield an estimate at the end of the window. The infectiousness is approximated by the distribution of the serial interval, which is defined as the time between the onset of symptoms of a case and the onset of symptoms of secondary cases infected by the primary case. Following previous work (Churches, 2020), we have set the distribution of serial intervals as a discrete gamma distribution with mean of 5 days and standard deviation of 3.4 days. Here, we concentrated on monitoring early stages of the epidemic in each country, when such simple exponential growth model is relatively accurate representation of the spread of the disease. Moreover, the estimated values are used mostly in the non-parametric Mann-Whitney test, which only considers their relative ordering, not exact values.

To avoid initial uncertainty in the reproductive number estimates due to small numbers of case reports, and to adjust for the differences in the start date of epidemics in each country, the seven-day interval for the first estimate (R100) starts on the day when 100 cases have been reported and the second estimate (R100+10) is taken 10 days later. The case incidence numbers have been smoothed over a window of 7 days in order to account for differences in testing procedures on different days of the week (i.e. no or little testing over the weekend in many countries). Such smoothing will not affect the parameters of exponential growth models. It has been verified that confidence intervals at chosen points of time are not unproportionally large.

### Application of Vazquez model

The number of infected individuals in the Vazquez model (Vazquez, 2006; Ziff and Ziff, 2020) has the form

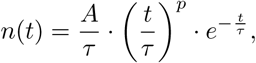

where *A, p*, and *τ* are parameters and *t* = 1 (units are days) corresponds to the first day of an infection. In practice, the available data does not report the number of infected individuals in the population due to limited testing availability and potential testing errors. Therefore we use the total number of active cases (confirmed - recovered - deaths) as a proxy for the total number of infected individuals. In countries with sufficient testing, we assume that the identified active cases represent a constant fraction of the total active cases and the formula for *n*(*t*) differs only in the constant factor *A*.

We used a nonlinear least squares method to infer the parameters in the above relationship from the data. However, instead of directly fitting the parameters *A*, *p*, and *τ*, we used an equivalent formulation

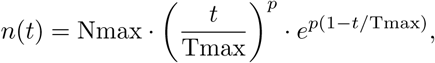

with parameters Nmax (the maximal number of active cases during the infection), *p* (the power of the polynomial growth term), and Tmax = *p* * *τ* (the time when the peak is reached). For consistency, we have truncated the data to reduce the impact of testing irregularities during the initial onset of epidemic. Therefore we start the data from the day when a certain number of active cases *N_a_* was reached. The threshold *N_a_* was chosen in proportion to the population in the country to reduce effects of randomness in reporting and to account for the spreading potential. Italy served as the reference with a threshold of 200 cases (threshold chosen was always at least 10).

## Data Availability

All datasets used for analysis in this study are publicly available in the repositories cited in the article.

## Acknowledgements

This work has been supported by the Slovak Research and Development Agency under the contracts no. APVV-18-0239 (T.V., B.B.), APVV-18-0308 (R.K.) and by the Scientific Grant Agency of the Slovak Republic under the grants no. 1/0458/18 (T.V.), 1/0463/20 (B.B.), 1/0755/19 (R.K.), and 1/0521/20 (K.B.)

